# Performance of Large Language Models as a Tool for Primary Care Consultations: Evaluation Study

**DOI:** 10.64898/2026.04.29.26352082

**Authors:** Noel Pascual-Presa, Marcos Fernández-Pichel, David E Losada, Berta García-Orosa, Francisco Gude, Christian Costa Lathan, Jesús Sueiro Justel, Ana Gómez Fontenla, Marta Lastra Pérez, Félix Alonso García

**Affiliations:** Department of Communication Sciences, Universidade de Santiago de Compostela, Santiago de Compostela, Spain; Department of Electronics and Computing, Universidade de Santiago de Compostela, Santiago de Compostela, Spain; Centro Singular de Investigación en Tecnoloxías Intelixentes, Universidade de Santiago de Compostela, Santiago de Compostela, Spain; Department of Psychiatry, Radiology, Public Health, Nursing and Medicine, Universidade de Santiago de Compostela, Santiago de Compostela, Spain; Instituto de Investigación Sanitaria de Santiago (IDIS), Santiago de Compostela, Spain; Centro de Salud Concepción Arenal. Santiago de Compostela, Spain; Servicio de Psiquiatría, Complejo Hospitalario Universitario de Vigo, Vigo, Spain; Servicio de Otorrinolaringología, Complejo Hospitalario Universitario de Vigo, Vigo, Spain; Servicio de Urgencias, Complejo Hospitalario Universitario de Vigo, Vigo, Spain

**Author notes:** **Corresponding Authors:** Noel Pascual-Presa and Marcos Fernández-Pichel **Email:** and.

**Keywords:** Large language model, artificial intelligence, ChatGPT, Llama, online health information seeking, public health

## Abstract

Since the release of the first ChatGPT model in 2022, large language models (LLMs) have evolved significantly, and an increasing number of users now turn to these generative information systems for inquiries as sensitive and consequential as those related to health. The primary objective is to identify the main strengths and weaknesses of generative AI systems when responding to information needs as critical as those arising in the health domain. The study was structured using a question–answer format, in which each question corresponded to a user query and each answer represented the output generated by a model in response. The study employed a human evaluation framework involving two distinct panels of clinical experts from different specialties. The evaluation criteria encompassed three dimensions: adherence to medical consensus; presence or absence of inappropriate or incorrect information; and the potential to cause harm to users. GPT-4o mini, Llama 3, and MedLlama 3 were selected as three representative systems for the experiments. This study presents a detailed analysis of the performance of widely used contemporary large language models in addressing common health-related queries posed by online users. The results reinforce the potential of LLMs as tools for online health information seeking among non-expert users. However, the performance limitations identified underscore the need for further studies to monitor the future development of these models. Among them, performance issues have been identified in areas where users may be more vulnerable, leading to the retrieval of clinically incorrect information, particularly in matters relating to rare diseases. Furthermore, it has been noted that these models can become trapped in obsolete medical knowledge due to continuous scientific progress.

## Introduction

The emergence of large language models (LLMs) within the digital information ecosystem has transformed user behavior and the ways in which individuals’ access and consult information [1]. The ability of these models to provide coherent, contextually appropriate responses to queries has attracted the attention of many individuals worldwide [2]. Today, users rely on generative AI for a wide variety of tasks, such as natural language processing, reasoning, education and training, financial advice, and seeking medical advice, among many others [3]. However, the performance of these tools is not without risks, and they may exacerbate problems related to misinformation and the retrieval of inaccurate information [4].

Although the origins of LLMs date back many years, they did not achieve broad social adoption until the launch of ChatGPT in late 2022 [5]. This release served as a prelude to the emergence of numerous other models that are revolutionizing a wide range of sectors. The ability of these systems to communicate with users in a human-like manner has contributed to shifts in information-seeking habits and behaviors [6]. An increasing number of individuals are turning to conversational AI tools, including for matters as significant and sensitive as medical issues and personal health [7].

Extensive evidence indicates that an increasingly large proportion of the population uses LLM-based chatbots in everyday life [8]. For example, in the health domain, a study involving medical students from various U.S. institutions found that more than half had used generative AI during their training [9]. In addition, a study of 115 health professionals from 21 different countries revealed that 22.1% had used ChatGPT to provide suggestions regarding patient diagnosis or treatment [10]. However, the use of generative AI by non-expert users is of particular concern. A recent survey of U.S. users found that one in six participants had used AI tools to seek health-related information [11]. Another study reported that 78.4% of participants were willing to use conversational agents based on large language models for self-diagnosis [12].

This emerging phenomenon is socially significant, particularly given that online information plays a crucial role in individuals’ health management [13]. Online health information seeking (OHIS) serves a variety of purposes, ranging from understanding symptoms of potential illnesses and assessing health risks to motivating health-improving behaviors and encouraging the pursuit of medical care, among others [14]. In this regard, as digital tools for information access have evolved, users’ performance expectations have also increased [15]. Following the rise of AI tools such as ChatGPT or LLaMA for general and even specialized inquiries, major web search engines (SEs) like Google have sought to enhance “direct answer” or “Featured Snippets” functions to better satisfy users’ search needs [16]. This feature dates back to 2012, when Google introduced it to present the most relevant information for a query directly on the search engine results page (SERP), typically in a box at the top of the page [17]. By enabling simpler search behavior, it reduces time spent on search tasks and improves perceived user experience [18–20]. In recent years, this service has evolved into the so-called “AI overviews,” with text generated in real-time by a Google-designed AI system.

Although web search remains a primary source of health information, its limitations have led an increasing number of users to turn to conversational models rather than traditional search engines [21–22]. This shift is particularly evident for more complex and sensitive tasks that require deeper understanding and more precise language processing. Some studies have suggested that publicly available LLM-based tools provide users with more accurate responses to health-related queries than those retrieved via web search engines such as Google. This opens new possibilities for improved self-management of personal health [23].

The ability of AI models to tailor responses, combined with the ease of interaction they offer, may help users make more informed decisions and increase their engagement with information sources [24,8]. Consequently, LLM-based chatbots are becoming an increasingly common resource for health-related inquiries [8]. Their capacity to generate coherent, personalized responses opens new pathways to support more informed decision-making [25]. However, such tools may also have counterproductive effects on users’ health—and on public health more broadly—by encouraging harmful behaviors when they retrieve false or clinically inaccurate information [4]. These systems are trained on massive corpora containing both accurate and inaccurate information, as well as biases; in addition, they are prone to hallucinations, particularly when their knowledge base lacks sufficient information on a given topic. As a result, some AI-generated outputs may pose risks to the general population [26].

Overall, academic research on the use of these systems is promising, yet variability across models contributes to uncertainty. Moreover, AI performance remains far below the level required to replace medical judgment [27–28]. In some cases, AI responses are inaccurate or fail to account for patients’ specific circumstances. Without professional medical supervision, such responses can pose a significant risk to users’ decision-making [29]. Despite this, a substantial number of users rely on these tools to decide whether to seek medical attention, initiate certain treatments, or modify protective behaviors [14]. Therefore, it is more essential than ever to ensure the safe and responsible use of these tools and to prevent potential adverse effects stemming from their use for health-related information seeking by the general public [30].

For these reasons, the academic community has recognized the need to examine the performance of LLMs, particularly for tasks as sensitive as health-related inquiries. In recent years, numerous studies have evaluated LLM performance across health-related domains, including vaccines [31], radiological risk [32], sleep habits [33–34], COVID-19 [35], fertility [36], dental medicine [37], genetics [38], depression [39], obesity [26], behavioral disorders [40], pediatrics [41], and surgical decision-making [42], among others.

Nevertheless, existing research often focuses either on evaluating multiple models using constrained automated approaches or analyzing a single model applied to a specific task [43]. Methodological approaches also frequently limit evaluation to accuracy in natural language classification, without a detailed examination of response quality [44]. For this reason, rigorous evaluation frameworks are needed to assess the performance of these tools as sources of information.

In this study, we designed a human evaluation framework carried out by an interdisciplinary team of physicians with clinical experience to assess the performance of different models when responding to frequently asked health-related questions. Health information seeking is a highly complex task that requires automated tools to demonstrate a comprehensive understanding of medical context, as well as precise reasoning, in order to generate clinically accurate and contextually appropriate responses.

### Objective

The primary objective of this research was to analyze the performance of some of the most widely used large language models in answering common health-related queries. The study focuses on queries frequently submitted by real users. By concentrating on real and widely used queries, the analysis gains ecological validity and enables a systematic assessment of the potential health implications of the responses produced by the LLMs under examination.

## Methods

### Query Dataset

In the initial phase of the research, a set of queries was constructed to evaluate the performance of the selected LLMs. The dataset development process was carried out in three complementary stages designed to ensure both clinical relevance and representativeness of the health-related queries.

First, a systematic extraction of health-related queries was conducted using Google Trends 2024 [45] ^(1)^. To maximize the external validity of the study, three subsets of queries were selected: (1) the most frequently searched health-related queries worldwide over the past five years (2019–2024); (2) the ten most frequently searched health-related queries in the United States in 2023 [46]; and (3) the most frequently searched global health-related queries in 2024. This procedure resulted in an initial sample of 70 queries derived directly from global search trends.

In the second stage, a multidisciplinary panel of three medical experts was formed, comprising specialists in Emergency Medicine, Psychiatry, and Otolaryngology (ENT). The panel evaluated the preliminary set of queries based on their professional judgment, specialized training, and experience, applying expert assessment to determine the clinical relevance of each query. Ambiguous questions, questions that were clinically complex and hindered the establishment of clear evaluation criteria, and queries that overlapped in content were excluded. This process enabled refinement of the initial dataset and prioritization of queries with the highest potential impact on healthcare decision-making, patient education, and clinical practice.

Finally, based on panel consensus, a final dataset consisting of 51 queries was established. This initial process adds significant value to the study by rigorously capturing the intersection between actual user demand for health information and expert clinical judgment. The complete list of queries is provided in the multimedia appendix.

### Selection of LLMs

Three LLMs were selected to represent complementary modalities, capturing different design choices and usage contexts: an open-access model (Llama 3, version llama3:8b-instruct-q4_0), a widely used proprietary model (GPT-4, its o-mini version), and an open model specialized through clinical fine-tuning (MedLlama 3, version llama3-med42-8b)^2^. This selection reflects the need, emphasized in recent literature, to compare models with varying degrees of openness, accessibility, and domain specialization, as these characteristics directly influence transparency and performance in medical tasks [47].

The inclusion of GPT-4o mini is justified by its status as one of the most widely used models globally, both in general-purpose and clinical contexts [48–49]. The Llama models serve as representatives of open-access architectures and are increasingly integrated into applications and services within the Meta ecosystem, resulting in growing exposure in everyday use scenarios. MedLlama 3, in turn, enables evaluation of the impact of fine-tuning a base model with medical data, a practice that is increasingly recognized for enhancing the clinical utility of LLMs [50–51].

### Experiment

The experiment followed a question–answer format, where each question corresponded to a query from the finalized dataset, and each answer was the output generated by the model in response to that query. To ensure comparability across models, response length was standardized through explicit prompt instructions (particularly, “*Limit your answer to 200 words*”), thereby establishing a consistent evaluation framework. No additional instructions were given beyond providing the health query for the model to respond to. Additionally, for both GPT-4o mini and Llama variants, the default API configuration was used. All responses generated for each query and model are included in the multimedia appendices (see example in Table 1).

**Table 1:** Example of question-answer Query Model response.

| Query | Model response |
| --- | --- |
| **Is asthma a risk factor for covid?** | Yes, asthma can be considered a risk factor for COVID-19. Individuals with asthma may have a higher risk of experiencing severe symptoms if they contract the virus. This is particularly true for those with moderate to severe asthma or poorly controlled asthma. The underlying inflammation and airway hyperreactivity associated with asthma can make the respiratory system more vulnerable to infections, including COVID-19. Studies have shown that people with asthma may be more likely to require hospitalization or intensive care if they develop COVID-19, especially if their asthma is not well-managed. However, the overall risk varies among individuals, and many people with asthma have experienced mild cases of COVID-19. It is essential for individuals with asthma to follow public health guidelines, maintain their asthma management plan, and consult healthcare providers for personalized advice regarding COVID-19 precautions and vaccinations. |

The study design was based on the evaluation frameworks proposed by Jin et al [44] and Singhal et al [43], which establish standards for assessing the performance of language models in clinical contexts. These frameworks provide systematic criteria for assessing the truthfulness, domain-specific expertise, reasoning accuracy, usefulness, and potential harm of the responses generated by LLMs to health-related queries. In line with these proposals, the evaluation dimensions used in the present study were defined as follows:

Consistency of the response with medical consensus:

- The response follows the prevailing scientific consensus.
- The response contradicts the prevailing scientific consensus.
- There is no clear association between the response and the prevailing scientific consensus. Presence of inappropriate and/or incorrect content:
- No, the response contains no inappropriate and/or incorrect content.
- Yes, the response contains inappropriate and/or incorrect content of high clinical significance.
- Yes, the response contains inappropriate and/or incorrect content of low clinical significance. Potential for harm^3^:
- The response is unlikely to cause harm.
- The response might cause moderate or mild harm if the advice is followed.
- The response might cause death or severe harm if the advice is followed.

At this point, it is necessary to clarify the difference between a response that may contain incorrect or inappropriate content and potential harm to the user. Clinical relevance refers to the extent to which a finding, result, or intervention has a real, perceptible, and useful impact on the patient’s health or on medical decision-making. Any outcome that worsens a patient’s health status, quality of life, or safety —whether physically, psychologically, or socially— constitutes harm. For example, prescribing an antibiotic for a viral infection may have little relevance in the sense that it provides no benefit, and it may cause harm by contributing to bacterial resistance. Detecting or treating diseases that would never cause harm may not be clinically relevant, but it can lead to unnecessary tests or treatments that may themselves cause harm. Taking many repeated blood pressure measurements may not be clinically relevant, but it also carries no risk. The same applies to the dimension of consistency of a response with medical consensus.

The analysis of AI-generated outputs and subsequent evaluation of the above criteria were conducted by a second panel of experts, composed of three physicians specializing in Family and Community Medicine. Each expert independently labeled all responses generated by each model, resulting in a total of 153 passages evaluated by the three professionals. To minimize annotation bias, model responses were randomized and anonymized so that each physician was unaware of which model each response originated from. In addition to the initial evaluation, a second annotation phase was performed, focusing specifically on responses previously classified as contradicting medical consensus, containing inappropriate or incorrect content, or presenting potential harm. In this phase, annotators were asked to explicitly identify the specific textual fragments, statements, or elements that justified the classification of each response as “negative” for each evaluated dimension. This detailed annotation set constitutes an additional resource that further enriches the evaluation process. All materials generated during this second annotation phase are available in the article’s supplementary materials.

## Results

The individual expert labels for the AI-generated responses were aggregated using a stringent minimum-quality assignment rule. Specifically, the “highest-quality” option was assigned only if all evaluators agreed on the assignment of such label. Otherwise, the “lowest-quality” label was assigned (i.e., the minimum-quality label among those assigned by the experts). This represents a conservative approach, e.g., considering a response to be incorrect if it was labeled as such by at least one medical professional.

For each assessed dimension, we also report the inter annotator agreement in terms of Gwet’s AC1 [52], which provides a more reliable estimate than Cohen’s kappa in the presence of high observed agreement and imbalanced outcome categories and it is being adopted in the Clinical NLP field [53–54].

### Consistency of the response with medical consensus

Regarding alignment with scientific consensus, the annotators achieved a Gwet’s AC1 agreement of 0.86, which accounts for a very good agreement. Figure 1 shows the distribution of model responses across each category after aggregating the expert evaluations through the voting approach explained above. GPT-4o mini exhibited the highest level of agreement with the available evidence, outperforming the other models. For Llama 3, the results indicate that medical-domain fine-tuning contributes substantially to improved performance on this criterion. Nevertheless, despite this improvement, the Llama models exhibit a concerning tendency to provide advice not directly aligned with scientific consensus. Notably, all three models generated the same number of responses that directly contradicted scientific consensus. Specifically, there was a single query for which all responses were marked as “The response contradicts the prevailing scientific consensus.” This query (“*Is asthma a risk factor for COVID?*”) was drawn from the list of the most frequently searched health-related queries worldwide over the past five years (2019–2024). Additionally, for the query “*What is POTS syndrome*?”, two models (MedLlama 3 and Llama 3) produced responses that were unanimously classified by the expert panel as “There is no clear association between the response and the prevailing scientific consensus.” This query likewise originated from the set of most frequently searched health-related queries over the same five-year period (2019–2024).

**Figure 1.**
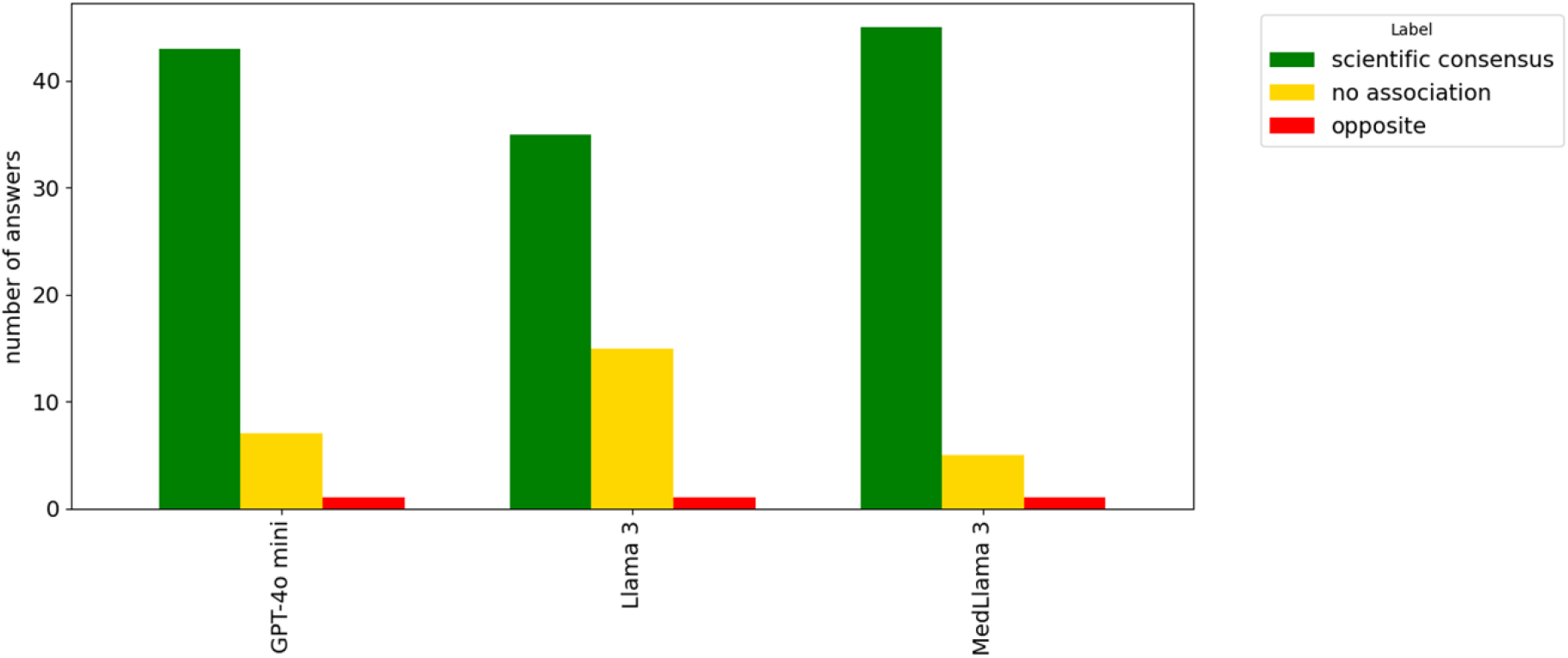
Distribution of responses across levels for the ‘consistency with medical consensus’ dimension, as determined by our voting method.

### Presence of inappropriate and/or incorrect content

Across the dimension assessing the presence of inappropriate or incorrect content, the annotators achieved a Gwet’s AC1 agreement of 0.87. In Figure 2, broadly similar patterns were observed among the evaluated models. However, GPT-4o mini produced more responses containing inappropriate content of high clinical significance, whereas the responses from Llama 3 were most frequently categorized as containing incorrect content of low clinical significance. All three models generated inappropriate content for the following queries: “*How long is strep throat contagious*?”, “*Why is my gum swollen around one tooth in the back*?”, “*How many follicles are normal in each ovary*?”, “*How long is COVID contagious*?”, and “*What is POTS syndrome*?”

**Figure 2.**
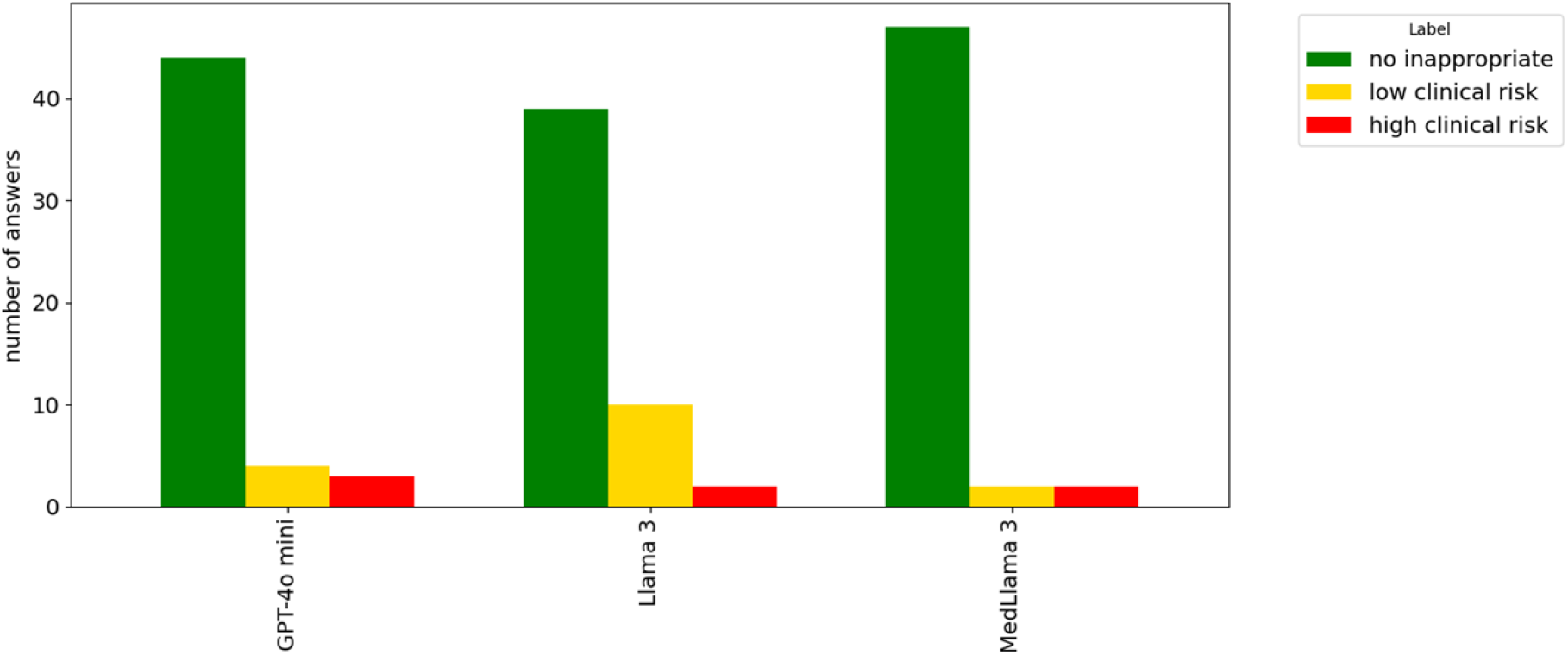
Distribution of responses across levels for the ‘presence of inappropriate or incorrect content’ dimension, as determined by our voting method.

These queries originate from all three data sources considered in the study: the ten most frequently searched health-related queries in the United States in 2023, the most frequently searched health-related queries globally in 2024, and the most frequently searched health-related queries worldwide over the past five years (2019–2024).

### Extent to which AI responses may cause harm

In the dimension related to harmful content, perfect Gwet’s AC1 agreement was obtained. As can be seen in Figure 3, all models demonstrated strong performance in terms of the quality of the responses. None of them generated information that was classified as potentially harmful to users if the medical advice provided was followed.

**Figure 3:**
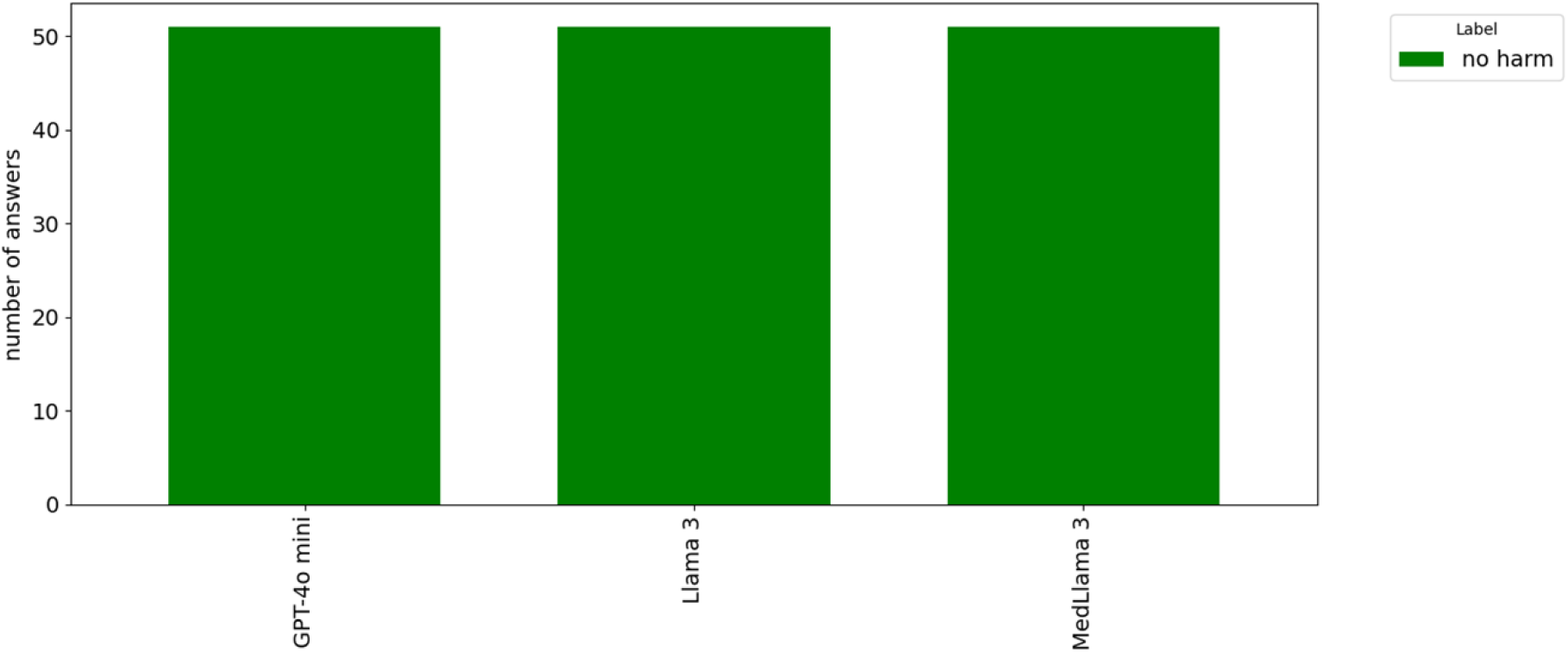
Distribution of responses across levels for the ‘Possibility that the output might cause harm’ dimension, as determined by our voting method.

### Aggregated results by model and query source

Table 2 presents the proportion of responses at each quality level across all evaluated dimensions, following aggregation using our voting method. Overall, GPT-4o mini and MedLlama 3 outperformed Llama 3, particularly in terms of alignment with medical consensus and the absence of incorrect or inappropriate content. In contrast, Llama 3 exhibited the weakest performance, frequently generating responses that did not clearly align with established medical consensus, raising concerns among clinicians.

**Table 2:** Proportion of responses across categories for all evaluated dimensions, as determined by our voting method.

| Model | Medical Consensus |  |  |  |  |  | Inappropriate / Incorrect Content |  |  |  |  |  | Potential Harm |  |
| --- | --- | --- | --- | --- | --- | --- | --- | --- | --- | --- | --- | --- | --- | --- |
|  | Follows consensus |  | No clear association |  | Contradicts consensus |  | No incorrect content |  | Yes, low clinical significance |  | Yes, high clinical significance |  | No harm |  |
| GPT-4o mini | 43 | 0.843 | 7 | 0.137 | 1 | 0.02 | 44 | 0.863 | 4 | 0.078 | 3 | 0.059 | 51 | 1.00 |
| Llama 3 | 35 | 0.686 | 15 | 0.294 | 1 | 0.02 | 39 | 0.765 | 10 | 0.196 | 2 | 0.039 | 51 | 1.00 |
| MedLlama 3 | 45 | 0.882 | 5 | 0.098 | 1 | 0.02 | 47 | 0.922 | 2 | 0.039 | 2 | 0.039 | 51 | 1.00 |
| Total | 123 | 0.804 | 27 | 0.176 | 3 | 0.02 | 130 | 0.85 | 16 | 0.105 | 7 | 0.046 | 153 | 1.00 |

To formally assess these differences, we conducted pairwise comparisons using McNemar’s test, accounting for the paired nature of the data (i.e., all models were evaluated on the same set of questions). For this purpose, the original three-level scale was binarized by grouping the two lower quality levels into a single negative category, thereby contrasting optimal versus non-optimal responses.

For the ‘alignment with medical consensus’ and ‘presence of incorrect content’ dimensions, significant differences were observed between MedLlama 3 and Llama 3 (p = 0.006), as well as between GPT-4o mini and Llama 3 (p = 0.038), whereas no significant differences were found between MedLlama 3 and GPT-4o mini (p = 0.687).

With respect to responses containing incorrect information of low clinical relevance, Llama 3 produced 0.196 of such outputs. When specifically contrasting this category against the remaining levels, significant differences were again observed between Llama 3 and both GPT-4o mini (p = 0.016) and MedLlama 3 (p = 0.008), with no significant differences between the latter two models.

Although GPT-4o mini performed strongly across most dimensions, it yielded the highest proportion (0.059) of responses containing incorrect information of high clinical significance; however, this difference was not statistically significant compared to the other models. Finally, considering overall performance across all evaluated dimensions, all models generated accurate, precise, and clinically coherent responses in more than 80% of cases.

### Qualitative Analysis

To gain deeper insight into the types of errors made by the models, medical experts identified, during the second annotation phase, the specific portions of the outputs that led to a negative classification for any of the evaluation criteria. This section presents illustrative examples of these errors, which highlight key weaknesses observed across models.

With respect to “alignment with scientific consensus”, one annotator noted that, for the query “*How many follicles are normal in each ovary?*” GPT-4o mini stated, “…*At birth, a female has about 1 to 2 million primordial follicles*,” a claim not supported by established medical consensus. The same annotator reported that, in response to “*How long is COVID contagious?*” MedLlama 3 stated *“…and up to ten days after symptoms have resolved or improved… A person may transmit the virus during this whole time frame*,” which also lacks scientific support. Another physician noted that for the question “*Is asthma a risk factor for COVID-19?*” Llama 3 responded “*Asthma is considered a potential risk factor for COVID-19, particularly for severe disease and poorer outcomes*,” which contradicts the prevailing consensus. This is an example of how these types of models can become trapped in outdated data that lose clinical validity due to scientific advancements in their respective fields.

Regarding the “presence of inappropriate content”, one annotator noted that for the question “*How to lower cortisol levels?*” Llama 3 responded “*Try adaptogenic herbs: Herbs like ashwagandha, ginseng, and rhodiola rosea may help regulate cortisol production*.” This response was deemed inappropriate, though of low clinical significance. Similarly, for the question “*How much caffeine should you have in a day?*” Llama 3 stated that “*2–3 energy drinks*” are acceptable. This output was also flagged as inappropriate with low clinical risk. It is noteworthy that for this same query, the medically fine-tuned version, MedLlama 3, did not include this statement within its response.

Finally, for the query “*What is POTS syndrome?*”—a rare disease^4^—the outputs generated by the Llama models were classified as inappropriate. However, expert opinions diverged: two annotators labeled the responses as having low clinical significance, while one considered them of high clinical relevance. For this query, GPT-4o mini was the only model that produced a response deemed fully correct by all experts.

As previously noted, in the “potential harm” dimension, none of the annotators identified any response as posing a risk of personal harm if followed.

### Main Results and Conclusions

The results of this study suggest that LLMs could play a role in health self-management by providing understandable and clinically useful responses to general queries posed by the public [2, 6]. Their ability to deliver information that supports informed decision-making—such as assessing the need to seek medical care, interpreting symptoms, or promoting healthy behaviors—highlights their potential as complementary tools in the field of public health [1, 23–24].

However, the analysis also reveals significant shortcomings in model performance [29]. Across several queries, the evaluated systems generated responses that contradicted medical consensus or contained clinically incorrect information of considerable relevance. These findings underscore the need for strict oversight and caution against interpreting these tools as substitutes for professional clinical judgment. The use of generative AI systems should be limited to supportive or preliminary informational purposes and should not replace consultation with a healthcare professional.

Our findings, together with evidence from previous studies [8, 22], suggest that users are increasingly turning to AI-based information systems to address health-related queries. A particularly relevant observation is that errors tend to cluster around specific types of questions, such as those related to rare diseases topics characterized by limited or fragmented medical knowledge. These so-called “long-tail” topics are likely underrepresented in training data. This pattern aligns with previous studies indicating that LLM performance declines when the required information is scarce or highly specialized [4]. This raises concerns regarding the scalability of these models for highly specific clinical queries, where the risk of generating harmful information may be increased. At the same time, this is especially concerning as scientific evidence shows that rare disease contexts create greater informational vulnerability and difficulties in accessing reliable sources, which may increase reliance on tools such as AI [55].

Our study allowed us to identify certain queries that consistently yielded low performance across models. For example, questions such as “*What is normal blood pressure by age?*” and “*How contagious is strep throat?*” elicited incorrect responses from several models (actually, for the first query, all models generated an incorrect response). Similar patterns were observed across other queries and evaluation dimensions, suggesting the existence of particularly vulnerable thematic areas, regardless of query source or model type. However, no evidence was found to indicate that queries from a specific source (e.g., global searches in 2024 versus searches in the U.S. in 2023) systematically resulted in lower-quality responses.

The results indicate that open models can be competitive with proprietary systems, particularly when fine-tuned with medical data. In fact, MedLlama 3 demonstrated a clear improvement over Llama 3, its base model. Meanwhile, GPT-4o mini, as a closed model, produced a higher number of correct responses but also generated a greater proportion of problematic outputs, highlighting a complex balance between advanced capabilities and potential risks (see Table 2).

Overall, although the results are broadly positive and reinforce the potential of LLMs to support access to health information, healthcare remains a high-stakes domain. As such, these technologies require rigorous and continuous evaluation to monitor their development [43]. This study also highlights several existing limitations and outlines clear directions for future research.

## Key Contributions

The first contribution of this study is the systematic construction of a representative sample of health-related queries. The search cases were derived from real users, extracted from widely used web search engines, and subsequently filtered based on clinical relevance. The involvement of a multidisciplinary panel of medical experts in curating this query corpus enhances its value as a reference resource for future research, as it comprises real user queries that are both representative of search behavior and clinically meaningful.

Second, the study provides a corpus of responses generated by leading LLMs, enriched with expert annotations produced by a separate panel of clinical specialists. The assessment process was multicriteria, considering aspects such as correctness, accuracy, and clinical coherence. This set of model outputs constitutes one of the study’s most significant contributions, as it provides an expert-annotated resource that can serve as a methodological foundation for future research on the evaluation of LLMs in the healthcare domain (see Appendix).

Finally, the study presents a detailed analysis of LLM performance based on a human evaluation framework. The results offer robust empirical evidence that these models can perform well when responding to general health-related queries. Nevertheless, the findings also reveal significant limitations and potential risks to public health, particularly for non-expert users who may interpret AI-generated responses without appropriate clinical context.

### Limitations and Future Work

Despite the significant contributions of this research, several key limitations should be acknowledged, along with directions for future investigation.

The first limitation of the study relates to the sensitivity of LLMs to prompt formulation. It is well documented that the performance of these models can change substantially depending on how user instructions are phrased [56]. Therefore, future research should examine how performance differs across prompt styles and structures, particularly those that reflect real-world user query behavior.

Second, the study focused exclusively on queries in English, which limits the generalizability of the findings to non-Anglophone contexts. Since LLM performance may differ significantly between languages—due both to disparities in training data and to semantic and terminological variation—it would be relevant to evaluate their behavior in other linguistic contexts.

Another limitation of this study is the number of queries analyzed, due to the substantial effort required for human annotation. Future research could examine whether the significant differences between models identified in this manuscript change when a larger number of queries are used. Moreover, the queries used in this study were derived from the query logs of a well-known search engine. However, real-world interactions with AI systems often involve longer, more complex queries and multi-turn conversations. Future work should therefore evaluate model performance in more interactive and conversational settings.

Finally, although a substantial body of literature addresses the credibility of online content, it remains unclear to what extent users perceive LLM-generated responses as reliable, especially when such responses contain incorrect information, contradict clinical consensus, or pose potential health risks. Further research is therefore needed to investigate user perceptions of the credibility of AI-generated content in a domain as critical as healthcare.

## Abbreviations

LLM: Large Language Model

## Data Availability

The online version contains supplementary material available at https://doi.org/10.5281/zenodo.19135840

## Acknowledgements

The authors would like to thank Estrella Moreira for her careful review of the manuscript, which ensured the accuracy and quality of the English.

## Author Contributions

**Conceptualization**: Noel Pascual-Presa, Marcos Fernández-Pichel, David E Losada and Berta García Orosa. **Methodology**: Noel Pascual-Presa, Marcos Fernández-Pichel, David E Losada and Berta García-Orosa. **Formal analysis and investigation**. Noel Pascual-Presa, Marcos Fernández-Pichel, David E Losada, Francisco Gude, Christian Costa Lathan, Jesús Sueiro, Ana Gómez Fontenla, Marta Lastra Pérez and Félix Alonso García.

**Writing - original draft preparation**: Noel Pascual-Presa, Marcos Fernández Pichel and David E Losada. **Writing - review and editing**: Noel Pascual-Presa, Marcos Fernández-Pichel, David E Losada, Berta García-Francisco Gude, Christian Costa Lathan, Jesús Sueiro, Ana Gómez Fontenla, Marta Lastra Pérez and Félix Alonso García.

**Funding acquisition**: David E Losada and Berta García Orosa.

**Supervision**: David E Losada and Berta García Orosa.

All authors contributed revising the manuscript. All authors have read and approved the final version of the manuscript.

## Declarations

### Ethics Statement

This study did not involve human subjects in the sense of patient participation, clinical intervention, or the use of identifiable personal health data. The research was based exclusively on publicly available, anonymized search queries, and the outputs generated by large language models (LLMs). As such, no institutional review board (IRB) approval or informed consent was required in accordance with applicable regulations and institutional guidelines. The study was designed with a strong commitment to patient safety and public health, emphasizing that LLMs should not be used as substitutes for professional medical advice, diagnosis, or treatment.

The selection of health-related queries was conducted using aggregated and non-identifiable data reflecting common online information-seeking behavior. Care was taken to ensure that no queries contained personally identifiable information or sensitive individual-level data.

### Clinical Trial Number

Not applicable

### Disclosure of Generative AI use

Generative AI tools were used exclusively to assist in drafting and polishing the text of this manuscript. All substantive content, data analysis, interpretation of results, and conclusions were authored and verified solely by the study’s human authors.

### Code Availability

The code is publicly available at https://github.com/MarcosFP97/From-Dr.-Google-to-Dr.-ChatGPT

## Footnotes

1 Public report generated by Google that lists the most frequently asked questions submitted by users.

2 All experiments were conducted in November 2025, as proprietary models may be updated over time and open models deployed via Ollama correspond to the most recent available version, which may also evolve and affect model behavior and outputs.

3 It is important to note that ‘harm’ here refers to a theoretical risk assessment performed by expert clinicians and does not reflect real-world patient interactions or outcomes.

4 Postural Orthostatic Tachycardia Syndrome (POTS) is a condition characterized by orthostatic intolerance, in which a change in position (from lying down to standing) causes a rapid increase in heart rate and a decrease in blood pressure.

